# Cardiotoxicity Associated with Targeted Therapies in Lung Cancer: A Retrospective Cohort Study of 2,427 Patients with FAERS Pharmacovigilance Validation

**DOI:** 10.1101/2025.11.13.25340140

**Authors:** Zhixuan Zhang

## Abstract

**Background:** Targeted cancer therapies have become a cornerstone of lung cancer treatment. However, their short- and long-term side-effects on the cardiovascular system in real-world remain poorly understood. Given that many early-stage cardiovascular injuries are clinically asymptomatic, large-scale dynamic serological monitoring in lung cancer patients undergoing these therapies may help clarify this issue.

**Methods:** We conducted a real-world study of 2,427 lung cancer patients treated with 82 targeted anticancer agents across seven therapeutic target classes, including epidermal growth factor receptor (EGFR), programmed cell death protein 1 (PD-1), vascular endothelial growth factor (VEGF), anaplastic lymphoma kinase (ALK), breakpoint cluster region-Abelson (BCR-ABL), human epidermal growth factor receptor 2 (HER2), and other targets, at Renji Hospital (2018–2024) and corroborated the findings using data from the FDA Adverse Event Reporting System (FAERS; 4,249 cardiac adverse events). Cardiotoxicity was defined according to the 2022 European Society of Cardiology (ESC) cardio-oncology guidelines as any new elevation in cardiac biomarkers following targeted anticancer therapy. Multivariable Cox regression models were used to assess cardiotoxicity risk across different drug targets. Nonlinear associations between PD-1 inhibitor exposure and cardiotoxicity were modeled using restricted cubic splines based on administration frequency and cumulative dose. Echocardiographic and quantitative ECG sensitivity analyses were performed to assess complementary cardiac functional and electrophysiologic patterns. Longitudinal biomarker trajectories were analyzed with generalized additive mixed models, and disproportionality signals were quantified using proportional reporting ratios (PRRs).

**Results:** 2,069 lung cancer patients with cardiac biomarker measurements were included, 326 (15.8%) developed cardiotoxicity. Among the targeted anticancer agents, PD-1 inhibitors showed association with the highest adjusted risk (hazard ratio (HR), 1.81; 95% confidence interval (CI), 1.43–2.28), followed by VEGF inhibitors (HR, 1.33; 95% CI, 1.06–1.67). The risk of PD-1 inhibitor-related cardiotoxicity followed a nonlinear exposure–response relationship, increasing sharply up to approximately four administrations, with a similar dose-based inflection at approximately 780 mg. Complementary sensitivity analyses showed a higher exploratory prevalence of reduced LVEF among VEGF-exposed patients and target-specific ECG patterns, including rate-related changes with VEGF-directed therapy and pre–post rate increases after PD-1-directed therapy. Longitudinal analysis revealed a marked divergence in the trajectories of B-type natriuretic peptide and creatine kinase–MB in PD-1-treated patients, whereas troponin levels remained largely stable. These findings were corroborated by FAERS analyses, in which PD-1 inhibitors exhibited the strongest disproportionality signals for severe cardiac adverse events (proportional reporting ratio (PRR), 1.84; 95% CI, 1.68–2.02), including immune-mediated myocarditis (PRR 28.41) and autoimmune myocarditis.

**Conclusions:** Across both a real-world clinical cohort and a large post-marketing pharmacovigilance dataset, PD-1 inhibitors were consistently associated with the highest cardiotoxicity burden, characterized by an early cycle-dependent rise in risk and distinct elevations in BNP and CK-MB. Combined hospital-based and FAERS evidence supports target-specific cardiac surveillance strategies during lung cancer targeted therapy.

## Introduction

Lung cancer remains the leading cause of cancer-related death worldwide (1). Over the past decade, substantial progress in treatment has been achieved through advances in targeted cancer therapies (1). Agents such as epidermal growth factor receptor (EGFR) tyrosine kinase inhibitors, vascular endothelial growth factor (VEGF) inhibitors, and immune checkpoint inhibitors particularly programmed death-1 (PD-1) antibodies has substantially improving progression-free and overall survival (2). However, as survival improves and treatment duration extends, cardiovascular toxicities have become increasingly recognized as important adverse effects that may offset therapeutic benefits and affect long-term outcomes (3).

Cardiotoxicity in the context of targeted therapy includes a wide spectrum of manifestations, ranging from asymptomatic elevations in cardiac biomarkers to clinically significant heart failure, arrhythmias, and vascular events (3). The underlying mechanisms are diverse, involving direct myocardial injury (4), endothelial dysfunction (5), and immune-mediated inflammation (6). Previous studies have reported that PD-1 inhibitors can trigger myocarditis, pericardial disease, and biomarker-defined myocardial injury (7), whereas VEGF inhibition may induce hypertension, thrombosis, and heart failure (8). Conversely, EGFR inhibitors have been associated with a relatively low incidence of cardiotoxicity, though cases of thromboembolic events, heart failure, cardiomyopathy, arrhythmia, and hypertension have been reported (9).

Nonetheless, most available data arise from small case series or clinical trial populations with stringent inclusion criteria, limiting their generalizability to real-world patients who often present with advanced age, comorbidities, and polypharmacy (10). Real-world evidence on the comparative cardiotoxic profiles of different targeted cancer therapies in lung cancer remains scarce (11). Furthermore, the cumulative risk associated with repeated drug administration is poorly defined (12). Given the growing use of targeted cancer therapies in clinical oncology, understanding the dose–response and temporal dynamics of cardiotoxicity is of immediate clinical relevance.

In our study, we first retrospective evaluated lung cancer cohort in Renji Hospital with 2,427 patients treated with 82 targeted cancer agents over a six-year follow-up. We aimed to quantify the incidence of cardiotoxicity, identify associated demographic and clinical risk factors, and compare the cardiotoxic risk among major targeted therapy classes. We explored nonlinear associations between drug exposure frequency and cardiotoxicity risk, modeled longitudinal trajectories of cardiac biomarkers to elucidate temporal patterns of cardiac response. To validate our findings, we incorporated post-marketing pharmacovigilance evidence from the FAERS database to characterize targeted-therapy–related cardiac adverse event patterns and class-specific cardiotoxicity. Our study provides a detailed, data-driven framework for individualized cardiac monitoring in targeted therapy recipients.

## Methods

### Single-center cohort analysis

#### Study Population

This retrospective cohort study included patients with lung malignancies treated at Renji Hospital between January 1, 2018, and June 26, 2024. Lung malignancy was identified using the International Classification of Diseases, 10^th^ Revision code C34. Eligible patients were required to have received at least one targeted anticancer agent during the study period. The study protocol was approved by the Ethics Committee of Renji Hospital in 2025 (approval No. 25Y12800500), and the requirement for written informed consent was waived because of the retrospective design and the use of de-identified clinical data.

After data cleaning, 2,427 eligible patients were identified (Figure 2). Patients without any available cardiac biomarker measurements, including creatine kinase-MB, B-type natriuretic peptide, N-terminal pro-B-type natriuretic peptide, troponin I, or high-sensitivity troponin I, were excluded (n = 358). The final analytical cohort therefore included 2,069 patients. Follow-up was defined as the time from initiation of targeted therapy to the first cardiotoxicity event or the end of follow-up, whichever occurred first.

**Figure 1.**
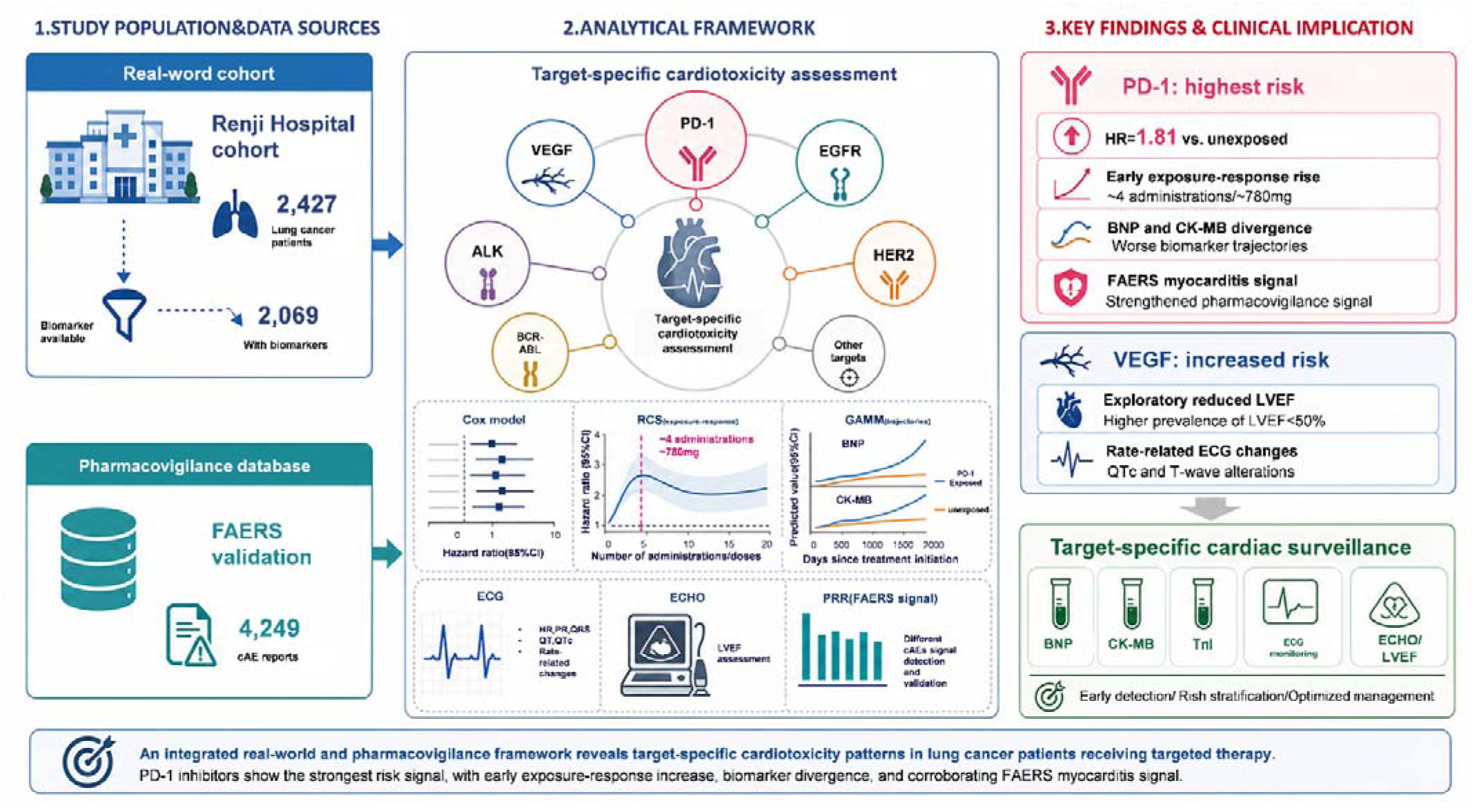
Central illustration.

**Figure 2.**
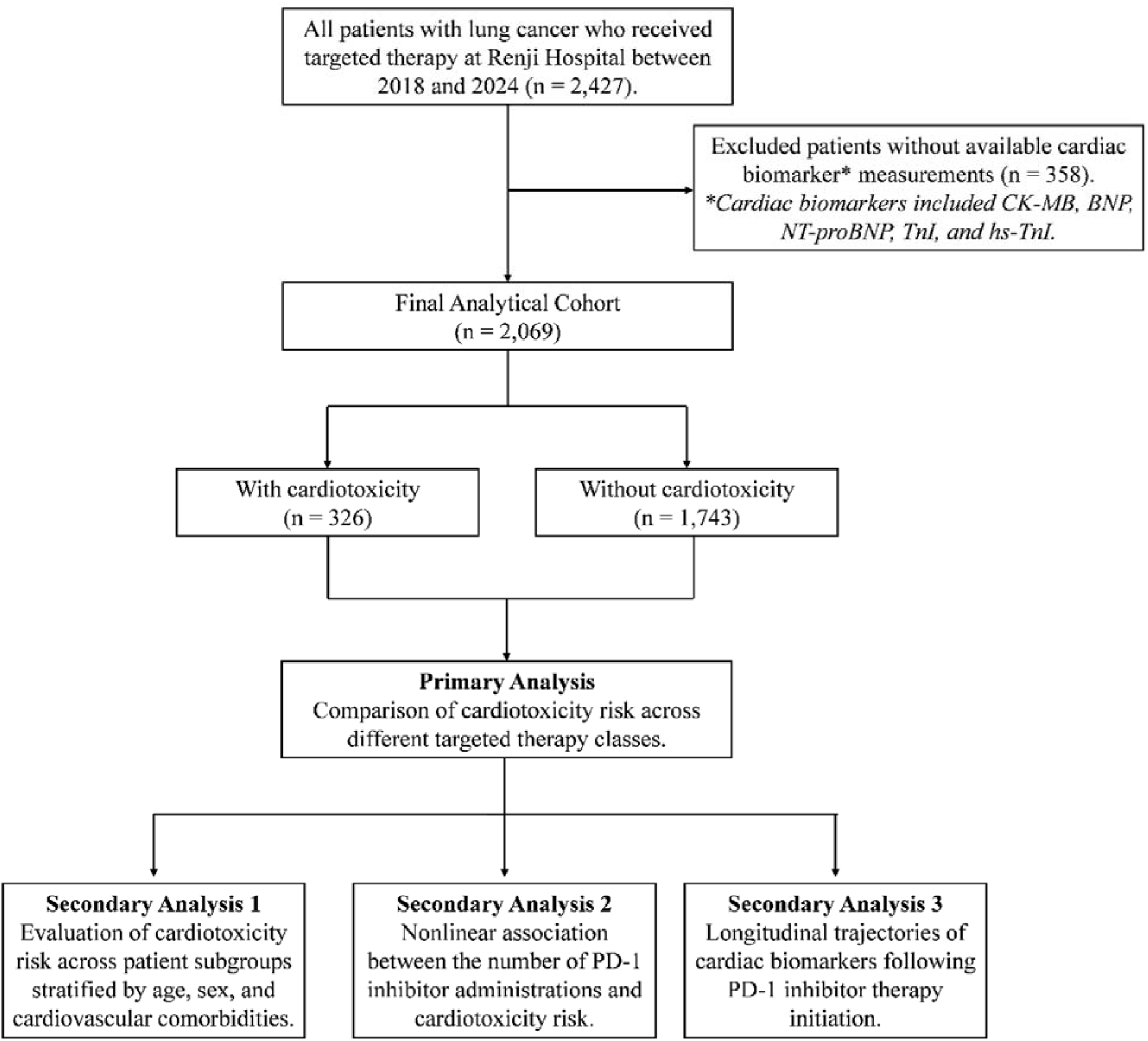
Study Flow and Analytical Framework (Renji Hospital cohort). Flowchart summarizing Renji Hospital cohort selection and analyses. Among 2,427 lung cancer patients treated with targeted therapies, 358 without cardiac biomarker data were excluded, yielding 2,069 for analysis. Patients were stratified into cardiotoxicity (n = 326) and non-cardiotoxicity (n = 1,743) groups. Primary analysis compared cardiotoxicity risk across targeted therapy classes. Secondary analyses included subgroup comparisons, nonlinear PD-1 exposure–response assessment, and longitudinal cardiac biomarker trajectory modeling. **Abbreviations: BNP, B-type natriuretic peptide; CK-MB, creatine kinase-MB; hs-TnI, high-sensitivity troponin I; PD-1, programmed cell death protein 1; TnI, troponin I.**

#### Statistical Analysis

The primary analysis compared the risk of cardiotoxicity across major targeted therapy classes, including EGFR inhibitors, PD-1 inhibitors, VEGF inhibitors, ALK inhibitors, and other agents, using multivariable Cox proportional hazards models adjusted for age, sex, and cardiovascular comorbidities. Secondary analyses included subgroup analyses according to demographic and clinical characteristics, assessment of nonlinear associations between PD-1 inhibitor exposure frequency and cardiotoxicity risk using restricted cubic spline models, and evaluation of longitudinal trajectories of cardiac biomarkers using generalized additive mixed models stratified by PD-1 inhibitor exposure.

#### Definition of Cardiotoxicity

Cardiotoxicity was defined as any new elevation in cardiac biomarkers—troponin I (> 0.04 ng/mL), high-sensitivity troponin (> 0.04 ng/mL), BNP (> 100 pg/mL), CK-MB (> 6.3 ng/mL), or NT-proBNP (> 20 pg/mL)—occurring on or after the first administration of a targeted drug. Cases were further classified based on the 2022 European Society of Cardiology (ESC) cardio-oncology guideline (13) into:(1) symptomatic CTRCD (heart failure requiring hospitalization or advanced therapy); (2) asymptomatic CTRCD (biomarker elevation only); (3) vascular toxicity; (4) hypertension; and(5) arrhythmia.

#### Drug Classification

Eighty-two targeted agents were grouped by their primary molecular targets (EGFR, PD-1, VEGF, ALK, BCR-ABL, HER2, and others; see Supplementary Table 1). Binary exposure variables (yes/no) were created for each target. Drug exposure frequency was calculated as the total number of administration orders documented by treating oncologists.

#### Covariates

Baseline variables included age, sex, and pre-existing cardiovascular and metabolic comorbidities diagnosed before initiation of targeted therapy. Comorbidities were identified using ICD-10 codes, and only the earliest diagnosis date was recorded for each condition. Specifically, hypertension was defined by codes I10–I13 and I15 (including hypertensive crisis I16 when applicable). Coronary artery disease and myocardial infarction were identified by I20 – I25. Arrhythmias and conduction disorders were defined by I44–I45, I47–I49. Heart failure and cardiomyopathy were captured by I50 and I42 (including drug-induced or secondary cardiomyopathy I42.7), with I51.7 (left ventricular hypertrophy) included when relevant. Thrombotic, atherosclerotic, and cerebrovascular disorders were identified using I26, I61–I64, I65– I67.2, I70 – I71, I80 – I83, I87.1 – I87.2, and I81 – I82, encompassing pulmonary embolism, stroke, peripheral arterial disease, and venous thromboembolism. Pulmonary hypertension was defined as I27.2, and valvular heart disease by I34–I37. Diabetes mellitus and dyslipidemia were identified using E10 – E14 and E78, respectively.

#### Single-center statistical analysis

In the single-center cohort from Renji Hospital, baseline characteristics were summarized according to the occurrence of cardiotoxicity. Continuous variables were compared using Student’s t test or the Wilcoxon rank-sum test, and categorical variables were compared using chi-square or Fisher’s exact tests, as appropriate.

The association between therapeutic target class and cardiotoxicity was evaluated using multivariable Cox proportional hazards models adjusted for age, sex, and baseline cardiovascular comorbidities. Follow-up was defined as the interval from treatment initiation to the first cardiotoxicity event or the end of follow-up, whichever occurred first. Hazard ratios with 95% confidence intervals were reported. Prespecified subgroup analyses were performed according to demographic and clinical characteristics.

Nonlinear exposure-response associations between PD-1 inhibitor exposure and cardiotoxicity were evaluated using restricted cubic spline functions in multivariable Cox models. Two PD-1 exposure metrics were examined: the number of PD-1 inhibitor administrations and the cumulative PD-1 inhibitor dose. We further performed echocardiographic sensitivity analyses based on available LVEF measurements and exploratory ECG analyses using linked quantitative ECG records. LVEF comparisons were conducted using Fisher’s exact tests. ECG parameters were analyzed using covariate-adjusted ordinary least-squares models with heteroscedasticity-consistent standard errors and paired pre-post comparisons using Wilcoxon signed-rank tests. Exact or nominal two-sided *P* values were reported for exploratory interpretation.

Longitudinal trajectories of cardiac biomarkers were analyzed using generalized additive mixed models to account for nonlinear temporal patterns and repeated measurements within patients. Four biomarkers with sufficient longitudinal data and clinical relevance for cardiotoxicity surveillance were included: BNP, CK-MB, myoglobin, and TnI. Biomarker concentrations were log-transformed as log(value + 1), and time since treatment initiation was modeled as a smooth function. For each biomarker, we compared a main-effect model with a shared temporal trajectory and an interaction model with PD-1 exposure-specific trajectories. Models included patient-level random intercepts and were adjusted for age, sex, and baseline cardiovascular comorbidities. Model fit was compared using likelihood ratio tests, Akaike information criterion, and marginal R². Improved fit of the interaction model was interpreted as evidence of divergent biomarker trajectories according to PD-1 inhibitor exposure.

### Validation analysis using FDA Adverse Event Reporting System (FAERS)

We obtained post-marketing safety data from the U.S. Food and Drug Administration (FDA) Adverse Event Reporting System (FAERS), a spontaneous reporting database that collects adverse event (AE) and medication error reports submitted by healthcare professionals, consumers, and pharmaceutical manufacturers (14). FAERS is structured around the FDA’s MedWatch reporting program and contains standardized information on patient demographics, drug exposures, indications, and clinical outcomes. Each submission in FAERS is recorded as an adverse event report (AER), which may contain one or multiple AEs for a given patient. Analyses in the present study were therefore performed at the report level (AER) after harmonizing drug names and consolidating multiple AEs within the same report. To ensure analytical validity, we followed FDA-recommended deduplication procedures and retained only the most recent version of each case.

Adverse events in FAERS are coded using Preferred Terms (PTs) from the Medical Dictionary for Regulatory Activities (MedDRA), an internationally standardized terminology system used by regulatory agencies and the biopharmaceutical industry. PTs represent distinct clinically meaningful adverse events and serve as the primary unit for safety signal evaluation.

### FAERS Cohort Identification and Analytical Procedures

We queried the FAERS database from 2004Q1 to 2023Q4 and identified 16,800,135 AERs. Reports involving non–targeted agents were excluded, resulting in 16,240,047 records. The specific agents included within each targeted therapy class are listed in Supplementary Table 2. A total of 560,088 reports containing any targeted therapy exposure were retained for further evaluation (Figure 6). Duplicate reports were removed by matching age, sex, country, indication, suspected drug, event date, and reaction, and only the most recent version was kept. Reports listing secondary suspect drugs and those involving patients aged ≤ 18 years were excluded, yielding 311,491 AERs. Subsequent restriction of indication resulted in 184,689 records, and limiting the cohort to lung cancer produced 63,908 reports.

After identifying 4,279 reports of targeted therapy–associated cardiac adverse events (cAEs) in lung cancer, we further excluded cases in which the treatment indication corresponded to primary or secondary cardiac neoplasms. These included “benign cardiac neoplasm”, “cardi ac neoplasm malignant”, “cardiac myxoma”, “pericarditis malignant”, “pericardial mesothelioma malignant”, “pericardial mesothelioma malignant recurrent”, “pericardial effusion malignant”, “metastases to heart”, “cardiac neoplasm unspecified”, “pericardial neoplasm”, “malignant pericardial neoplasm”, “benign pericardium neoplasm”, “cardiac fibroma”, “cardiac haemangioma benign”, “cardiac neurofibroma”, “cardiac teratoma”, “cardiac valve fibroelastoma”, “primary cardiac lymphoma”, “leukemic cardiac infiltration”, “pericardial lipoma”, and “cardiac lipoma”. A total of 30 reports met these exclusion criteria, resulting in a final cohort of 4,249 cardiac adverse event reports.

The final analytical cohort consisted of 4,249 reports documenting cAEs. Three analyses were conducted: (1) comparison of cAEs incidence across targeted therapy classes; (2) estimation of proportional reporting ratios (PRRs) for severe cAEs (PRR ≥ 3); and (3) heatmap-based profiling of PRRs for severe cAEs (PRR ≥ 3) associated with individual targeted therapies.

### Definition of cAEs

For this study, all cardiac adverse events in the overall frequency analysis (Figure 7A) were identified based on MedDRA PT codes within the cardiac disorders system organ class.

For the disproportionality analysis (Figure 7B) and the heatmap analysis (Figure 7C), we focused on severe cAEs with documented clinical significance. A total of 25 specific severe cAEs were included: pericardial effusion, myocarditis, cardiac tamponade, immune-mediated myocarditis, atrioventricular block complete, autoimmune myocarditis, pericarditis constrictive, pericardial disease, arrhythmia supraventricular, tachycardia paroxysmal, immune-mediated pericarditis, atrial flutter, cardiac failure acute, cardiac dysfunction, cardiopulmonary failure, long QT syndrome, bradycardia, pericarditis, sinus bradycardia, cardiac fibrillation, acute coronary syndrome, sinus tachycardia, cardiomyopathy, supraventricular tachycardia, and left ventricular dysfunction.

These 25 events were selected based on demonstrating a PRR ≥ 3.0 in at least one of the six drug target categories (HER2, EGFR, PD-1/PD-L1, ALK, VEGF, or PD-1 inhibitors), indicating a strong disproportionality signal and clinically meaningful association. Events with fewer than 3 reported cases for a specific drug target were excluded from PRR calculation for that target and designated as “NA” (not available) in the heatmap to ensure statistical reliability.

### Descriptive characterization of targeted therapy–related cAEs

We conducted a descriptive analysis of cAEs associated with targeted therapies in lung cancer in the FAERS database. Clinical and reporting characteristics were summarized across six major targeted drug classes (PD-1, PD-L1, EGFR, HER2, ALK, and VEGF inhibitors). Variables extracted included patient age, sex, and time to onset of cAEs, which was calculated as the interval between therapy initiation and event start date. Annual reporting frequencies were tabulated to reflect temporal adoption patterns of each therapeutic class. Continuous variables are presented as median values with interquartile ranges, and categorical variables are reported as counts and percentages.

### Disproportionality analysis

Disproportionality analysis was performed using the proportional reporting ratio (PRR) to assess the association between drug targets and severe cAEs (15,16). PRR and 95% CI was calculated as:

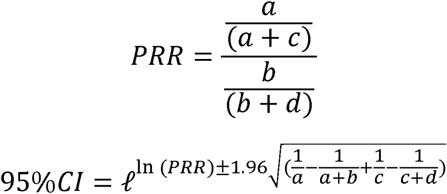

where *a* represents the number of reports with the specific cAE for the drug target of interest, *b* represents the total number of reports for that drug target, *c* represents the number of reports with the specific cAE for all other drugs, and *d* represents the total number of reports for all other drugs.

A cAE signal was considered valid when the number of reported cases was at least three and the lower bound of the 95% CI of the PRR exceeded 1.0, indicating a strong association with the corresponding targeted therapy. For each PT, PRR estimates and their 95% CIs were calculated, and statistical significance was further assessed using chi-square tests, with *P* < 0.01 regarded as significant. Only PTs with at least three reports were retained for the heatmap analysis to ensure signal stability and data reliability.

### Fatal outcome definition and analysis

Fatal outcomes were evaluated in the final FAERS cohort of targeted therapy–related cardiac adverse event reports among patients with lung cancer. Formal fatal outcome status was defined using the FAERS OUTC table, with a report classified as fatal if any linked outcome code (outc_cod), after standardization to lower case and trimming of whitespace, was equal to “death”. Death-related reaction terms were not used as the primary definition of fatal outcome. The denominator was the target-class–expanded set of cAE reports, consistent with the target-specific FAERS analyses; therefore, a report with overlapping exposure to more than one target class could contribute to more than one target-class category.

For each target class (PD-1, PD-L1, EGFR, HER2, ALK, and VEGF inhibitors), we calculated the number and proportion of cAE target-report combinations with formal FAERS death outcome. To further characterize the clinical phenotype of fatal cardiovascular AE reporting, MedDRA preferred terms (PTs) recorded in reports with death outcome were grouped into cardiovascular PT categories: cardiac/respiratory arrest, heart failure/shock, ischaemic events, myocarditis, pericardial disease, arrhythmia/conduction, and other cardiovascular PTs. Because a single FAERS report may include multiple reaction PTs, the PT-composition analysis was performed at the PT-report record level and was interpreted as a reporting-pattern analysis rather than an adjudicated analysis of cardiovascular cause of death.

## Results

### Study Framework

Figure 1 summarizes the integrated study framework, linking the Renji Hospital real-world cohort with FAERS pharmacovigilance validation. This design enabled target-specific evaluation of cardiotoxicity through clinical risk modeling, PD-1 exposure–response analysis, longitudinal biomarker trajectories, ECG and echocardiographic sensitivity analyses, and external disproportionality assessment. Together, these complementary analyses highlighted PD-1 inhibitors as the most consistent cardiotoxicity signal across both clinical and pharmacovigilance data.

### Baseline Characteristics

Cardiotoxicity was defined as a new elevation in any of the 4 cardiac biomarkers (BNP, NT-proBNP, TnI, hs-TnI) following initiation of targeted therapy. The cohort was stratified into patients with cardiotoxicity (n = 326, 15.8%) and without cardiotoxicity (n = 1,743, 84.2%). All baseline demographic, clinical, and treatment characteristics for both groups are summarized in Table 1. Of the 2,069 patients included in the analysis, 326 (15.8%) developed cardiotoxicity after targeted therapy. The median follow-up period was 527 days (IQR 192–974). The mean age did not differ significantly between groups (*P* = 0.096). Cardiotoxicity was more common in males (67.8% vs. 61.6%, *P* = 0.038).

**Table 1.**
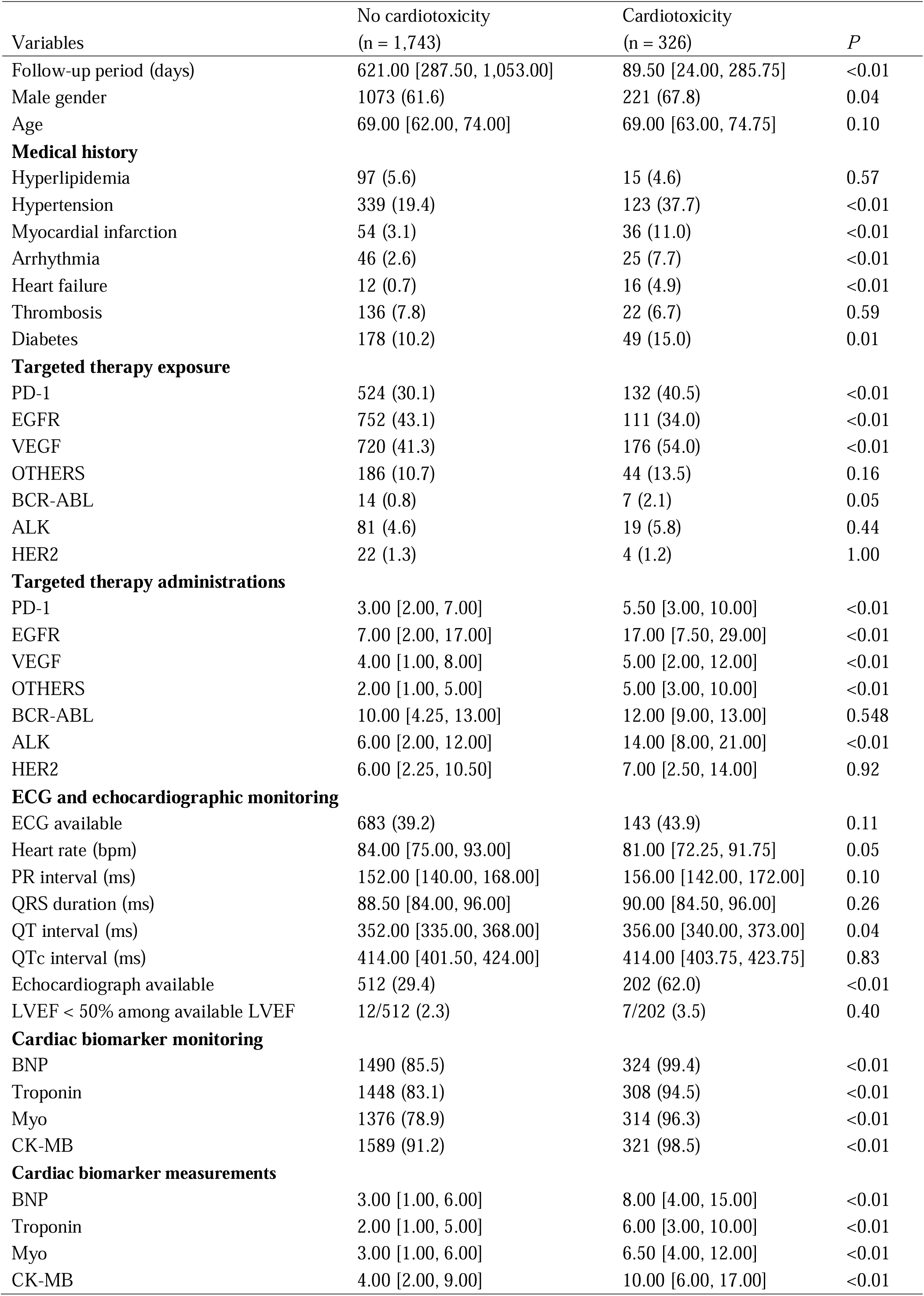
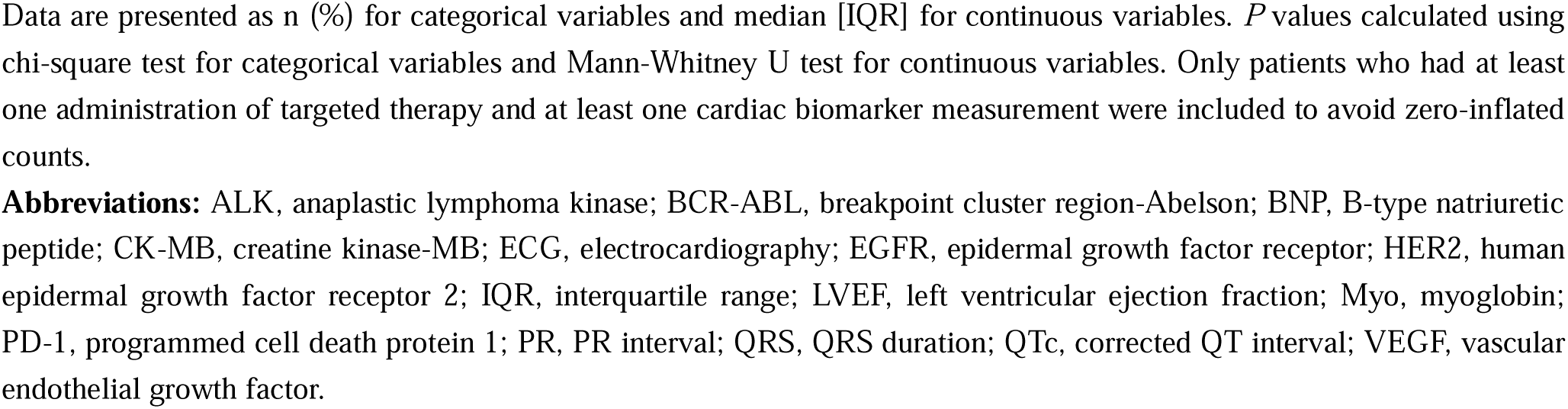
Baseline characteristics.

| Variables | No cardiotoxicity<br>(n = 1,743) | Cardiotoxicity<br>(n = 326) | *P* |
| --- | --- | --- | --- |
| Follow-up period (days) | 621.00 [287.50, 1,053.00] | 89.50 [24.00, 285.75] | <0.01 |
| Male gender | 1073 (61.6) | 221 (67.8) | 0.04 |
| Age | 69.00 [62.00, 74.00] | 69.00 [63.00, 74.75] | 0.10 |
| **Medical history** |  |  |  |
| Hyperlipidemia | 97 (5.6) | 15 (4.6) | 0.57 |
| Hypertension | 339 (19.4) | 123 (37.7) | <0.01 |
| Myocardial infarction | 54 (3.1) | 36 (11.0) | <0.01 |
| Arrhythmia | 46 (2.6) | 25 (7.7) | <0.01 |
| Heart failure | 12 (0.7) | 16 (4.9) | <0.01 |
| Thrombosis | 136 (7.8) | 22 (6.7) | 0.59 |
| Diabetes | 178 (10.2) | 49 (15.0) | 0.01 |
| **Targeted therapy exposure** |  |  |  |
| PD-1 | 524 (30.1) | 132 (40.5) | <0.01 |
| EGFR | 752 (43.1) | 111 (34.0) | <0.01 |
| VEGF | 720 (41.3) | 176 (54.0) | <0.01 |
| OTHERS | 186 (10.7) | 44 (13.5) | 0.16 |
| BCR-ABL | 14 (0.8) | 7 (2.1) | 0.05 |
| ALK | 81 (4.6) | 19 (5.8) | 0.44 |
| HER2 | 22 (1.3) | 4 (1.2) | 1.00 |
| **Targeted therapy administrations** |  |  |  |
| PD-1 | 3.00 [2.00, 7.00] | 5.50 [3.00, 10.00] | <0.01 |
| EGFR | 7.00 [2.00, 17.00] | 17.00 [7.50, 29.00] | <0.01 |
| VEGF | 4.00 [1.00, 8.00] | 5.00 [2.00, 12.00] | <0.01 |
| OTHERS | 2.00 [1.00, 5.00] | 5.00 [3.00, 10.00] | <0.01 |
| BCR-ABL | 10.00 [4.25, 13.00] | 12.00 [9.00, 13.00] | 0.548 |
| ALK | 6.00 [2.00, 12.00] | 14.00 [8.00, 21.00] | <0.01 |
| HER2 | 6.00 [2.25, 10.50] | 7.00 [2.50, 14.00] | 0.92 |
| **ECG and echocardiographic monitoring** |  |  |  |
| ECG available | 683 (39.2) | 143 (43.9) | 0.11 |
| Heart rate (bpm) | 84.00 [75.00, 93.00] | 81.00 [72.25, 91.75] | 0.05 |
| PR interval (ms) | 152.00 [140.00, 168.00] | 156.00 [142.00, 172.00] | 0.10 |
| QRS duration (ms) | 88.50 [84.00, 96.00] | 90.00 [84.50, 96.00] | 0.26 |
| QT interval (ms) | 352.00 [335.00, 368.00] | 356.00 [340.00, 373.00] | 0.04 |
| QTc interval (ms) | 414.00 [401.50, 424.00] | 414.00 [403.75, 423.75] | 0.83 |
| Echocardiograph available | 512 (29.4) | 202 (62.0) | <0.01 |
| LVEF < 50% among available LVEF | 12/512 (2.3) | 7/202 (3.5) | 0.40 |
| **Cardiac biomarker monitoring** |  |  |  |
| BNP | 1490 (85.5) | 324 (99.4) | <0.01 |
| Troponin | 1448 (83.1) | 308 (94.5) | <0.01 |
| Myo | 1376 (78.9) | 314 (96.3) | <0.01 |
| CK-MB | 1589 (91.2) | 321 (98.5) | <0.01 |
| **Cardiac biomarker measurements** |  |  |  |
| BNP | 3.00 [1.00, 6.00] | 8.00 [4.00, 15.00] | <0.01 |
| Troponin | 2.00 [1.00, 5.00] | 6.00 [3.00, 10.00] | <0.01 |
| Myo | 3.00 [1.00, 6.00] | 6.50 [4.00, 12.00] | <0.01 |
| CK-MB | 4.00 [2.00, 9.00] | 10.00 [6.00, 17.00] | <0.01 |

Data are presented as n (%) for categorical variables and median [IQR] for continuous variables. *P* values calculated using chi-square test for categorical variables and Mann-Whitney U test for continuous variables. Only patients who had at least one administration of targeted therapy and at least one cardiac biomarker measurement were included to avoid zero-inflated counts.
**Abbreviations:** ALK, anaplastic lymphoma kinase; BCR-ABL, breakpoint cluster region-Abelson; BNP, B-type natriuretic peptide; CK-MB, creatine kinase-MB; ECG, electrocardiography; EGFR, epidermal growth factor receptor; HER2, human epidermal growth factor receptor 2; IQR, interquartile range; LVEF, left ventricular ejection fraction; Myo, myoglobin; PD-1, programmed cell death protein 1; PR, PR interval; QRS, QRS duration; QTc, corrected QT interval; VEGF, vascular endothelial growth factor.

Pre-existing cardiovascular comorbidities were significantly more prevalent among patients who developed cardiotoxicity, including hypertension (37.7% vs. 19.4%, *P* < 0.001), myocardial infarction (11.0% vs. 3.1%, *P* < 0.001), heart failure (4.9% vs. 0.7%, *P* < 0.001), and diabetes (15.0% vs. 10.2%, *P* = 0.014). No significant differences were observed in the baseline prevalence of hyperlipidemia or thrombosis. In terms of oncological treatment, patients in the cardiotoxicity group were more likely to receive PD-1 inhibitors (40.5% vs. 30.1%, *P* < 0.001) and VEGF inhibitors (54.0% vs. 41.3%, *P* < 0.001), whereas EGFR inhibitor use was lower (34.0% vs. 43.1%, *P* = 0.003).

The cardiotoxicity group also received a higher median number of treatment administrations, including PD-1 inhibitors (5.5 vs. 3.0, *P* < 0.001), EGFR inhibitors (17.0 vs. 7.0, *P* < 0.001), VEGF inhibitors (5.0 vs. 4.0, *P* = 0.002), ALK inhibitors (14.0 vs. 6.0, *P* = 0.002), and other targeted agents (5.0 vs. 2.0, *P* < 0.001).

In terms of ECG and echocardiographic monitoring, ECG records were available for 683 patients without cardiotoxicity (39.2%) and 143 patients with cardiotoxicity (43.9%), with no significant between-group difference (*P* = 0.11). Among patients with available ECG data, heart rate was numerically lower in the cardiotoxicity group (81.00 [72.25, 91.75] vs 84.00 [75.00, 93.00] bpm; *P* = 0.05), whereas the QT interval was slightly longer (356.00 [340.00, 373.00] vs 352.00 [335.00, 368.00] ms; *P* = 0.04). PR interval, QRS duration, and QTc interval were comparable between groups. Echocardiographic data were more frequently available in patients with cardiotoxicity than in those without cardiotoxicity (62.0% vs 29.4%; *P* < 0.01). Among patients with available LVEF measurements, the prevalence of LVEF < 50% did not differ significantly between groups (3.5% vs 2.3%; *P* = 0.40).

Monitoring of cardiac biomarkers was significantly more frequent in patients who developed cardiotoxicity (all *P* < 0.001). Testing rates were higher for BNP (99.4% vs. 85.5%), CK-MB (98.5% vs. 91.2%), Myoglobin (96.3% vs. 78.9%), and Troponin (94.5% vs. 83.1%), with similar trends observed for hs-TnT (43.6% vs. 19.5%) and NT-proBNP (39.3% vs. 19.9%).

Among those tested, the median number of biomarker assessments was also significantly higher in the cardiotoxicity group (all *P* < 0.003), particularly for CK-MB (10.0 [IQR 6.0–17.0] vs. 4.0 [IQR 2.0–9.0], *P* < 0.001), BNP (8.0 [IQR 4.0–15.0] vs. 3.0 [IQR 1.0–6.0], *P* < 0.001), hs-TnT (8.0 [IQR 4.0–16.8] vs. 4.0 [IQR 2.0–9.0], *P* < 0.001), Troponin (6.0 [IQR 3.0–10.0] vs. 2.0 [IQR 1.0–5.0], *P* < 0.001), and Myoglobin (6.5 [IQR 4.0–12.0] vs. 3.0 [IQR 1.0–6.0], *P* < 0.001).

### Drug Class Associations

Multivariable Cox regression analysis was performed to evaluate the association between specific targeted therapies and the risk of cardiotoxicity, adjusting for age, gender, and baseline cardiovascular comorbidities (including hypertension, myocardial infarction, arrhythmia, heart failure, thrombosis, diabetes, and hyperlipidemia) (Figure 3A).

**Figure 3.**
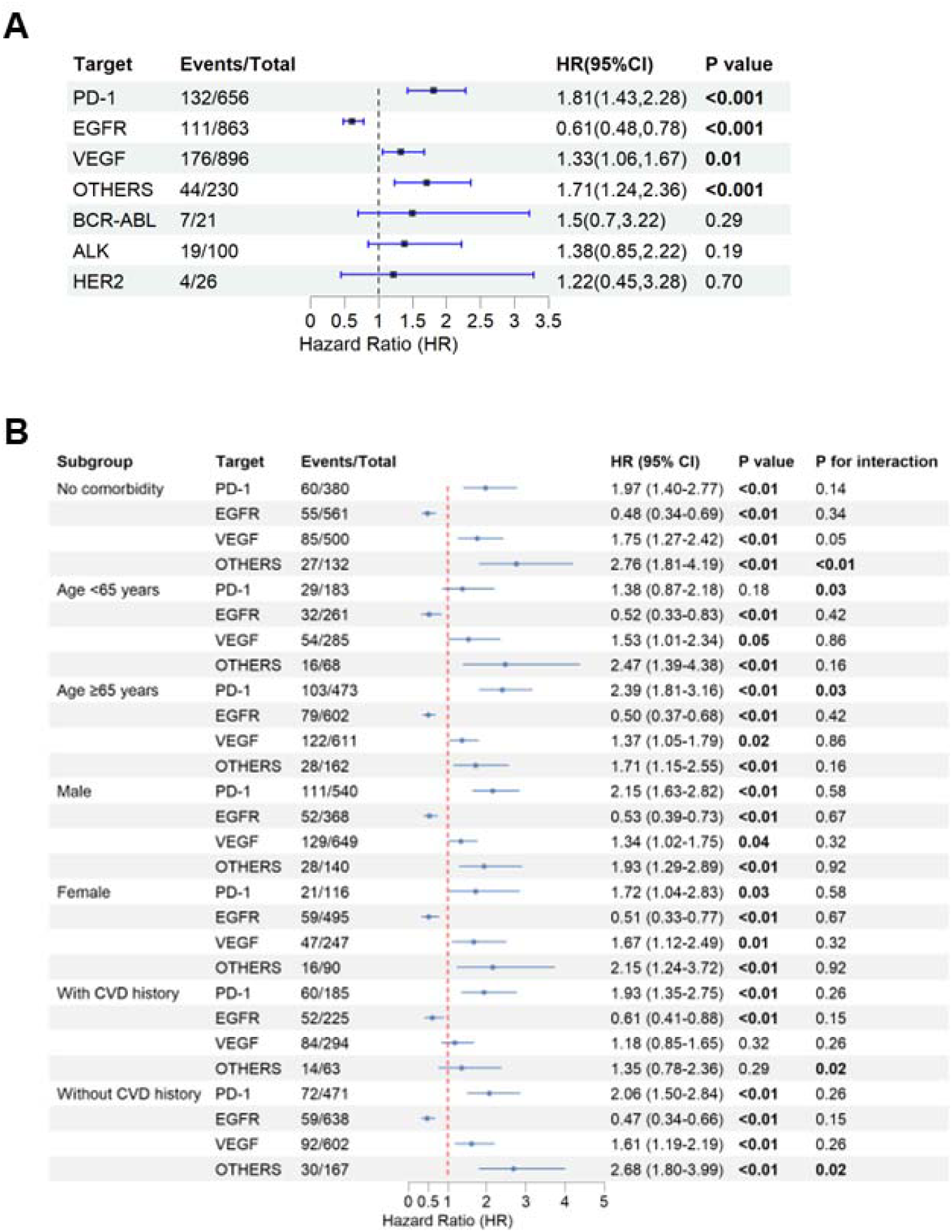
Cardiovascular Risk Associated with Different Targeted Therapies Overall and by Patient Subgroups. **(A) shows overall hazard ratios (HRs) and 95% confidence intervals (CIs) for cardiovascular events associated with different targeted therapies. (B) presents subgroup analyses stratified by baseline comorbidities, age, sex, and cardiovascular disease (CVD) history. All analyses were adjusted for age, sex, baseline CVD, diabetes, hypertension, hyperlipidemia, and cumulative treatment cycles using multivariable Cox proportional hazards models. The vertical dashed line represents HR = 1.0 (no difference in risk). *P* for interaction tests whether treatment effects differ significantly across subgroups.** **Abbreviations**: PD-1, programmed cell death protein 1; EGFR, epidermal growth factor receptor; VEGF, vascular endothelial growth factor; BCR-ABL, breakpoint cluster region-Abelson; ALK, anaplastic lymphoma kinase; HER2, human epidermal growth factor receptor 2; CVD, cardiovascular disease.

After adjustment, PD-1 inhibitor use was associated with the highest risk of cardiotoxicity (HR = 1.81, 95% CI, 1.43–2.28), followed by Other targeted therapy (HR = 1.71, 95% CI, 1.24–2.36), VEGF inhibitors (HR = 1.33, 95% CI, 1.06–1.67), and ALK inhibitors (HR = 1.38, 95% CI, 0.85–2.22). In contrast, EGFR inhibitor use was associated with a significantly lower risk of cardiotoxicity (HR = 0.61, 95% CI, 0.48–0.78).

Subgroup analysis revealed heterogeneous treatment effects across different targeted therapies and patient characteristics (Figure 3B). For PD-1 inhibitors, increased cardiovascular risk was consistently observed across most subgroups (all *P* < 0.01), except in patients aged < 65 years (HR = 1.38, 95% CI, 0.87–2.18, *P* = 0.18). A significant interaction was detected between age and PD-1 therapy (*P* for interaction = 0.03), with older patients (≥ 65 years) demonstrating higher risk (HR = 2.39, 95% CI, 1.81–3.16) compared to younger patients.

In contrast, EGFR inhibitors consistently showed protective cardiovascular effects across all subgroups examined, with hazard ratios ranging from 0.47 to 0.61 (all *P* < 0.01), without significant effect modification by any patient characteristics (all *P* for interaction > 0.05).

VEGF inhibitors demonstrated increased cardiovascular risk in most subgroups, though the effect was not significant among patients with pre-existing cardiovascular disease (CVD) history (HR = 1.18, 95% CI, 0.85–1.65, *P* = 0.32). For other targeted agents, a significant interaction with comorbidity burden was observed (*P* for interaction < 0.01), with the highest risk noted in patients without comorbidities (HR = 2.76, 95% CI, 1.81–4.19, *P* < 0.01). Additionally, cardiovascular history significantly modified the effect of other targeted therapies (*P* for interaction = 0.02), with attenuated risk observed in patients with pre-existing CVD (HR = 1.35, 95% CI, 0.78–2.36, *P* = 0.29) compared to those without CVD history (HR = 2.68, 95% CI, 1.80–3.99, *P* < 0.01).

### Exposure-response association of PD-1 inhibitors and complementary cardiac sensitivity analyses

Restricted cubic spline analysis showed a nonlinear association between PD-1 inhibitor exposure and cardiotoxicity risk (Figure 4A; *P* for nonlinearity = 0.002). Risk increased rapidly up to approximately four administrations (HR, 2.00; 95% CI, 1.50–2.65) and then increased more gradually at higher exposure counts. A similar pattern was observed when exposure was modeled as cumulative PD-1 inhibitor dose (overall *P* < 0.001; *P* for nonlinearity < 0.001; Figure 4B), with an estimated turning point at 781.6 mg, displayed as 780 mg. At this dose, the adjusted HR was 2.75 (95% CI, 1.98–3.82) relative to non-exposure.

**Figure 4.**
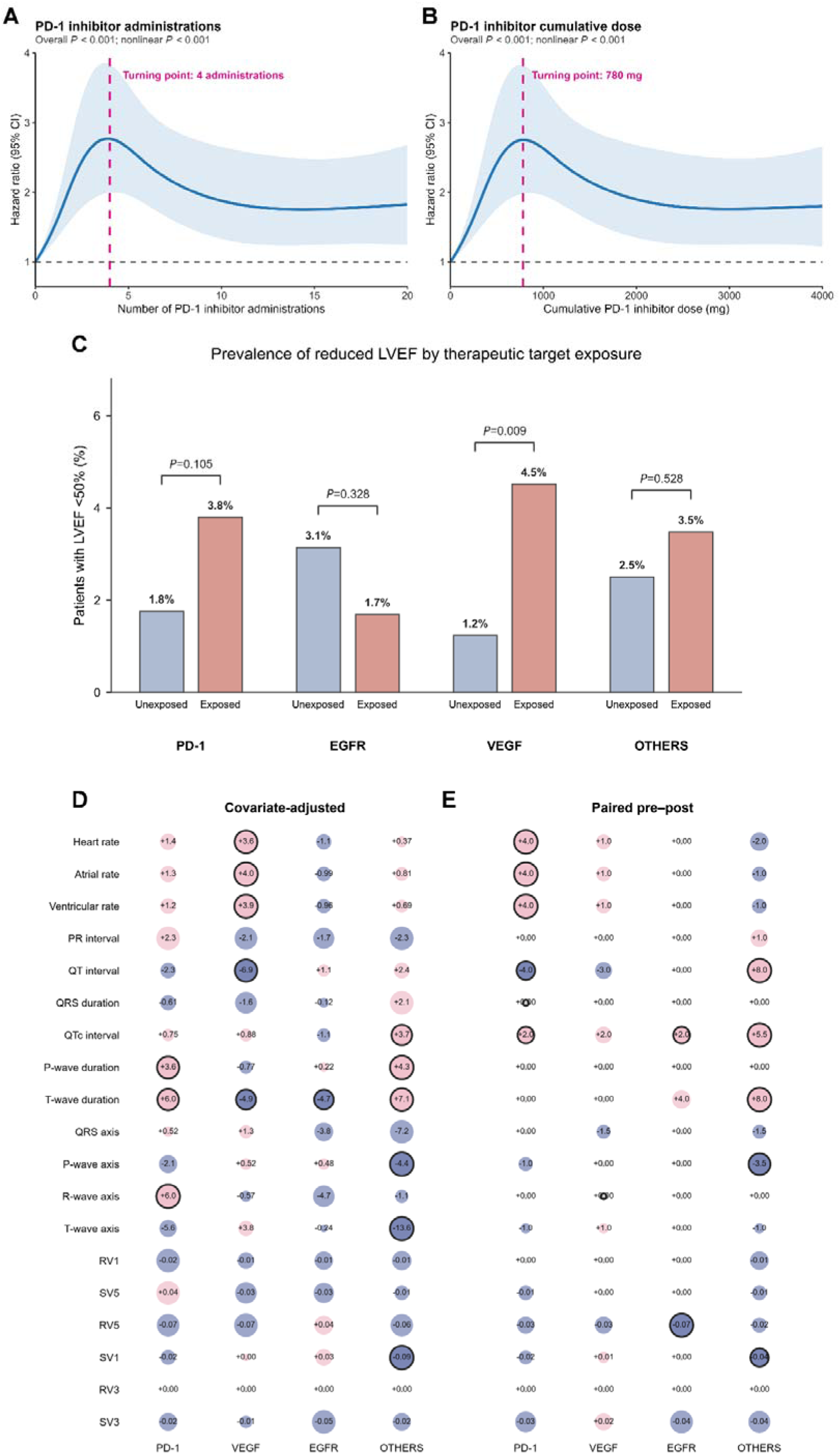
Exposure-response, echocardiographic, and electrocardiographic sensitivity analyses in the Renji Hospital cohort. (A) Restricted cubic spline analysis showing the nonlinear association between the number of PD-1 inhibitor administrations and cardiotoxicity risk. The solid blue line represents the adjusted hazard ratio, and the shaded area indicates the 95% confidence interval. The dashed horizontal line indicates HR = 1. The vertical dashed line marks the estimated turning point at four administrations. (B) Restricted cubic spline analysis showing the association between cumulative PD-1 inhibitor dose and cardiotoxicity risk. Cumulative dose was calculated from documented administration records using drug-specific per-administration doses. The estimated turning point was 780 mg. (C) Echocardiographic sensitivity analysis comparing the prevalence of reduced LVEF, defined as minimum recorded LVEF < 50%, between exposed and unexposed patients within each therapeutic target category. Target-specific exposure groups were defined independently and were not mutually exclusive. *P* values were calculated using two-sided Fisher’s exact tests. (D) Exploratory patient-level ECG analysis of 19 quantitative ECG parameters among patients linked to ECG records. For each patient, valid ECG measurements were summarized using the median value for each parameter. Circle size and color indicate the magnitude and direction of covariate-adjusted mean differences between exposed and unexposed patients for each target category. (E) Paired pre-post ECG analysis comparing ECG measurements before and after initiation of target-directed therapy. The last ECG within 365 days before treatment and the first ECG within 365 days after treatment were selected for each patient. Within-patient changes were evaluated using two-sided Wilcoxon signed-rank tests. Pink indicates an increase, blue indicates a decrease, and black outlines indicate nominal *P* < 0.05. **Abbreviations:** bpm, beats per minute; CI, confidence interval; ECG, electrocardiography; EGFR, epidermal growth factor receptor; HR, hazard ratio; LVEF, left ventricular ejection fraction; PD-1, programmed cell death protein 1; PR, PR interval; QRS, QRS duration; QTc, corrected QT interval; RCS, restricted cubic spline; VEGF, vascular endothelial growth factor.

In the echocardiographic sensitivity analysis, the prevalence of reduced LVEF differed numerically across target-specific exposure groups (Figure 4C). Reduced LVEF was more frequent among patients exposed to VEGF-directed therapy than among unexposed patients (4.5% [14/310] vs 1.2% [5/404]; *P* = 0.009). The corresponding differences were not statistically significant for PD-1-directed therapy (3.8% vs 1.8%; *P* = 0.105), EGFR-directed therapy (1.7% vs 3.1%; *P* = 0.328), or other targets (3.5% vs 2.5%; *P* = 0.528). Given the descriptive nature of this sensitivity analysis, the VEGF-associated difference should be interpreted as exploratory.

Exploratory ECG analyses identified target-specific electrophysiologic patterns (Figure 4D and 4E). In covariate-adjusted analyses, VEGF-directed therapy showed the most coherent rate-related pattern, with higher heart rate, atrial rate, and ventricular rate, as well as a shorter QT interval. PD-1-directed therapy was associated with longer T-wave and P-wave durations. In paired pre-post analyses, PD-1-directed therapy was associated with median increases of 4 bpm in heart rate, atrial rate, and ventricular rate. Other nominal ECG findings were observed across target groups, but these results should be interpreted as exploratory signals requiring confirmation in dedicated analyses with prespecified ECG endpoints.

### Temporal Trajectories of Cardiac Biomarkers by PD-1 Exposure

Figure 5 shows the modeled longitudinal trajectories of four cardiac biomarkers according to PD-1 inhibitor exposure over a follow-up of up to 2500 days after treatment initiation. All generalized additive models converged with satisfactory fit.

**Figure 5.**
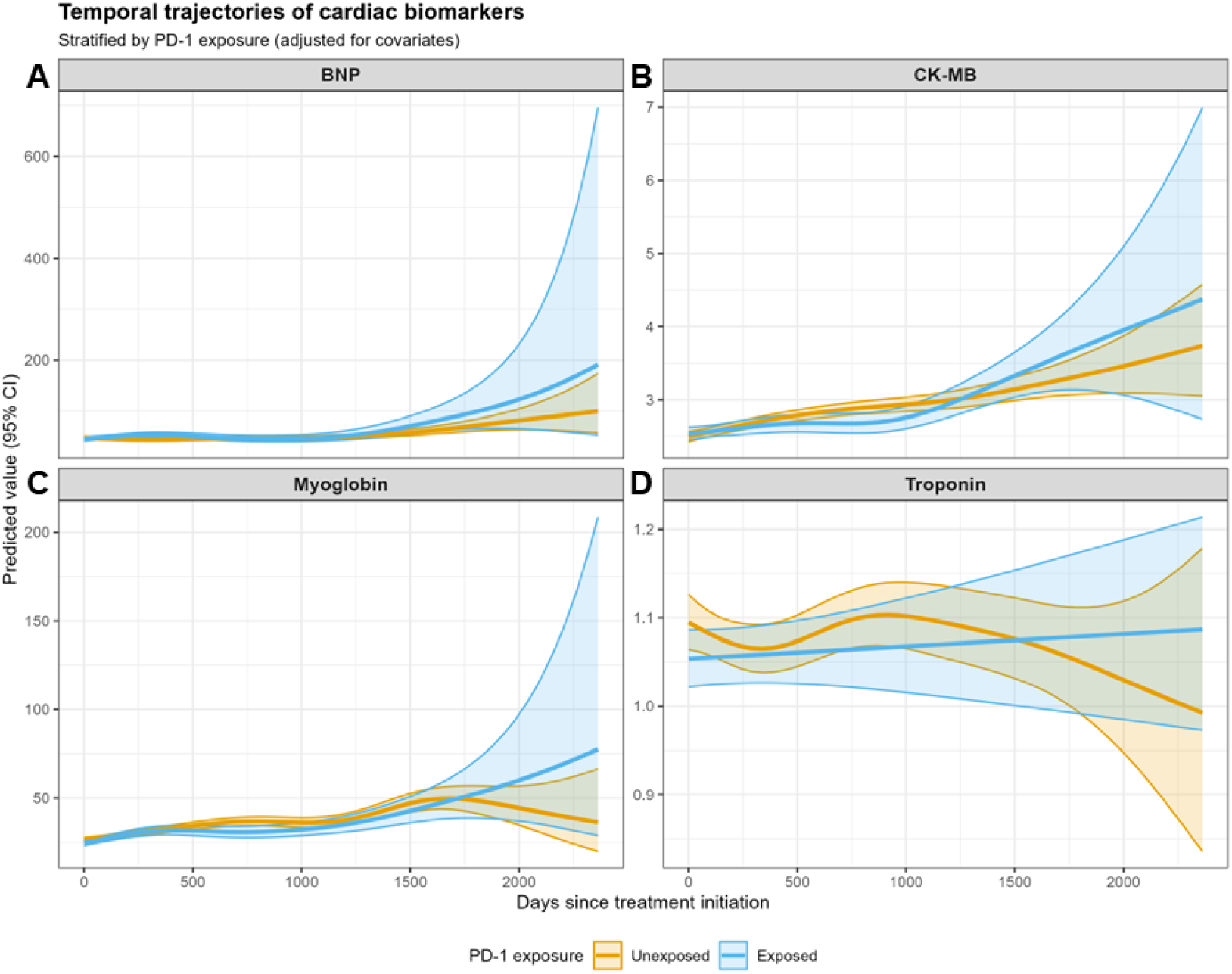
Temporal trajectories of cardiac biomarkers according to PD-1 inhibitor exposure. Predicted longitudinal trajectories of cardiac biomarkers over 2,500 days after treatment initiation are shown for (A) BNP, (B) CK-MB, (C) myoglobin, and (D) troponin. Trajectories were estimated using generalized additive mixed models adjusted for age, sex, PD-1 administration frequency, and baseline cardiovascular comorbidities. Blue lines indicate patients exposed to PD-1 inhibitors, and orange lines indicate unexposed patients. Shaded areas represent 95% confidence intervals. **Abbreviations: BNP, B-type natriuretic peptide; CI, confidence interval; CK-MB, creatine kinase-MB; PD-1,programmed cell death protein 1.**

For BNP, PD-1–exposed patients displayed a distinct late-phase rise, with both groups remaining stable around 50 pg/mL during the first 1500 days, after which the exposed group showed an exponential increase exceeding 200 pg/mL by day 2500, whereas the unexposed group maintained levels around 50–100 pg/mL. CK-MB levels increased gradually in both groups, with a steeper slope in the PD-1–exposed group emerging by 1000 days and becoming more pronounced after 1500 days, reaching 4 ng/mL by day 2500 versus 3.5 ng/mL in the unexposed group. Myoglobin followed a similar pattern to BNP, remaining stable until about 1500 days, then rising sharply in the exposed group to over 75 ng/mL by day 2500, compared with decreases to around 25 ng/mL in the unexposed group. In contrast, troponin trajectories were largely comparable between groups, showing only minor fluctuations (1.05 to 1.10 ng/mL in exposed; 1.10 to 1.00 ng/mL in unexposed) with overlapping confidence intervals throughout the follow-up. Overall, BNP and myoglobin exhibited the most pronounced delayed elevations in the PD-1–exposed cohort, suggesting potential cumulative cardiotoxic effects associated with prolonged PD-1 inhibitor exposure, whereas CK-MB showed moderate sensitivity and troponin remained largely stable.

### Comparison of temporal trajectories between PD-1 **–** treated and untreated groups

Temporal trajectories of cardiac biomarkers were compared between PD-1–exposed and unexposed patients using generalized additive mixed models including smooth time-by-exposure interaction terms (Table 2).

**Table 2.** Model comparison results assessing the association between PD-1 inhibitor exposure and longitudinal trajectories of four cardiac biomarkers. Δ*AIC* represents the reduction in Akaike Information Criterion when including the PD-1 × time interaction term, indicating model improvement. *R² (interaction)* and *R² (main)* denote the proportion of variance explained by the interaction and main-effect models, respectively. *No. of observations* indicates total biomarker measurements; *No. of patients with cardiotoxicity* refers to individuals who developed cardiotoxicity and contributed at least one biomarker record. **Abbreviations: BNP, B-type natriuretic peptide; CK-MB, creatine kinase-MB; ΔAIC, delta Akaike information criterion; R^2^, marginal coefficient of determination**.

| Biomarker | No. of observations | No. of patients with<br>cardiotoxicity | ΔAIC | *P*-value | $R^2$<br>(interaction) | $R^2$<br>(main) |
| --- | --- | --- | --- | --- | --- | --- |
| BNP | 6934 | 1399 | 10.36 | **0.002** | 0.131 | 0.128 |
| CK-MB | 9799 | 1574 | 5.04 | **0.0192** | 0.042 | 0.041 |
| Myoglobin | 6414 | 1308 | 1.88 | 0.0641 | 0.096 | 0.095 |
| Troponin | 3589 | 1094 | 1.08 | 0.0896 | 0.013 | 0.012 |

Model comparison by likelihood ratio tests revealed significant heterogeneity in temporal patterns for two biomarkers (Table 2). BNP showed the strongest exposure-related divergence (ΔAIC = 10.36, *P* = 0.002; R2(interaction) = 0.131 vs R2(main) = 0.128; 6,934 observations from 1,399 patients), followed by CK-MB (Δ AIC = 5.04, *P* = 0.019; R2(interaction) = 0.042 vs R2(main) = 0.041; 9,799 observations from 1,574 patients). In contrast, Myoglobin and TnI showed comparable temporal trajectories regardless of PD-1 exposure after adjustment for covariates.

The marginal increase in R^2^ values between the interaction and main-effect models indicates that incorporating time-by-exposure interactions modestly improved model fit, particularly for BNP, reflecting a distinct exposure-dependent evolution over time.

### Baseline characteristics of targeted therapy–related cAEs in FAERS

A total of 4,249 adverse event reports involving targeted therapies and cardiac adverse events were identified in patients with lung cancer. Table 3 summarizes the demographic and reporting characteristics across drug classes. Median age varied across therapies, ranging from 64 years in ALK and VEGF inhibitor reports to 71 years among EGFR inhibitor reports. Sex distribution differed substantially by target: EGFR, HER2, and ALK inhibitors were associated with a predominance of female reports, whereas PD-1 and PD-L1 inhibitors were predominantly reported in males. Time to onset of cAEs also varied, with the longest delays observed for VEGF (median 60 days) and EGFR inhibitors (median 41 days), and the shortest for HER2 inhibitors (median 15 days). Reporting-year distributions demonstrated clear temporal adoption patterns across drug classes. PD-L1 inhibitors showed no reports before 2016 and demonstrated rapid growth from 2019 onward, with annual counts increasing from 77 in 2019 to 93 in 2020, followed by 82 in 2021, 99 in 2022, and 128 in 2023. In contrast, EGFR-related cAEs were reported continuously from 2004 through 2023, beginning with 47 –37 –32 reports in 2004 –2006 and remaining consistently represented in later years (e.g., 104 in 2017, 115 in 2019, 105 in 2020). VEGF inhibitors likewise exhibited steady reporting throughout the entire period, with early contributions in 2004–2007 (2–13 reports) and persistent signals in later years (e.g., 21 in 2017, 25 in 2022, 29 in 2023).

**Table 3.**
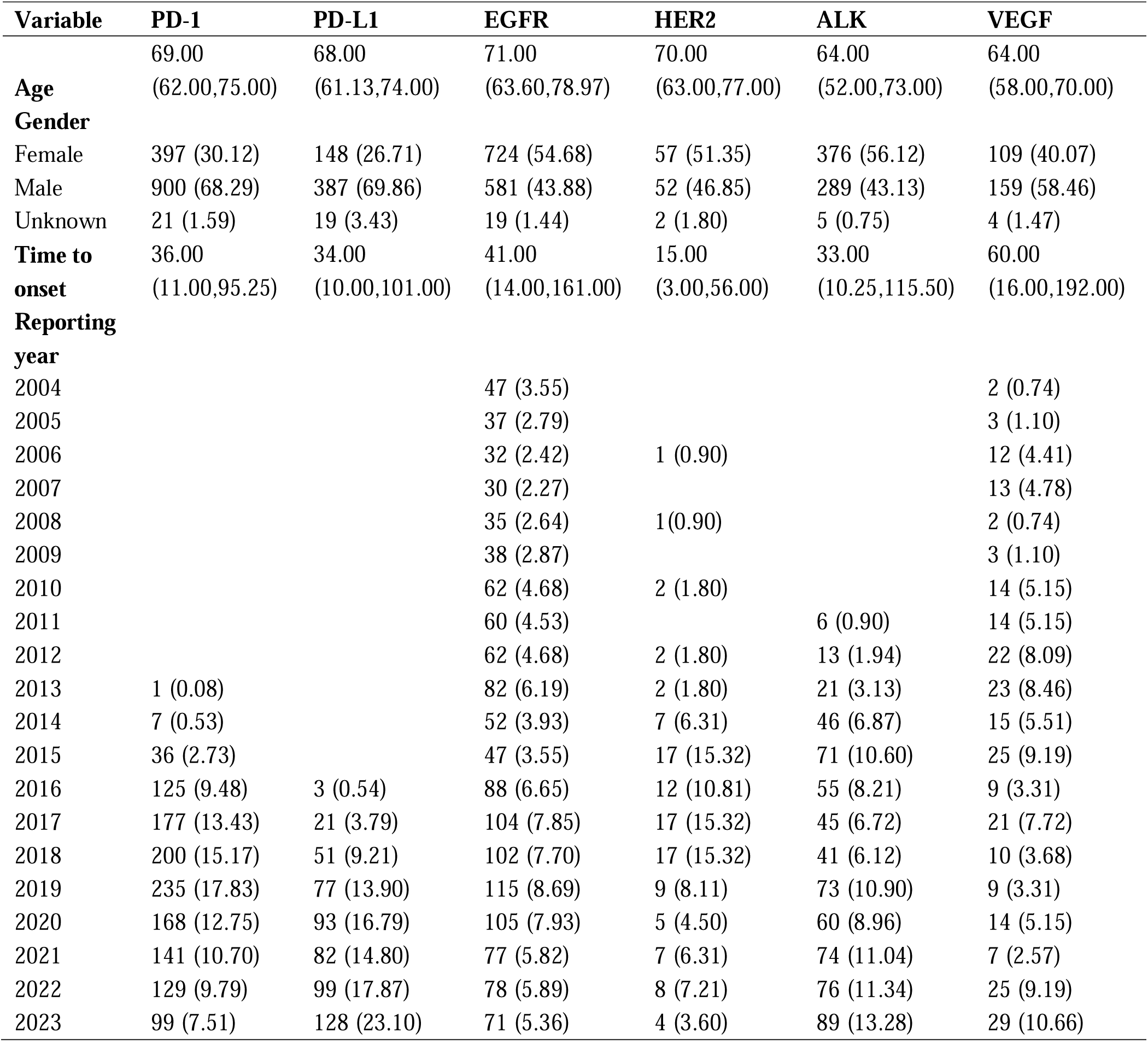
Characteristics of cardiac adverse event reports associated with targeted therapies in lung cancer (FAERS database, 2004–2023) Continuous variables are summarized as median [interquartile range], and categorical variables are expressed as counts with corresponding percentages (n [%]). **Abbreviations**: AER, adverse event report; ALK, anaplastic lymphoma kinase; cAE, cardiac adverse event; EGFR, epidermal growth factor receptor; FAERS, FDA Adverse Event Reporting System; HER2, human epidermal growth factor receptor 2; PD-1, programmed cell death protein 1; PD-L1, programmed death-ligand 1; PRR, proportional reporting ratio; VEGF, vascular endothelial growth factor.

### Targeted data processing and construction of the final analytical cohort

From a total of 16,800,135 reports in the FAERS database (2004Q1–2023Q4), 560,088 entries involved targeted therapies after excluding all non–targeted agents. Following removal of reports without lung cancer as the primary indication, 63,908 lung-cancer–related targeted-therapy reports were retained. After deduplication based on demographics, indication, suspected drug, event date, and reaction, and after excluding secondary suspect drugs and patients aged ≤ 18 years, 311,491 unique reports were retained. Subsequently, 59,629 reports without any cardiac adverse events (cAEs) were excluded. Among the 4,279 reports containing cAEs, an additional 30 cases with cardiac tumor–related indications (see Methods for detailed indication definitions) were removed. The final analytical cohort therefore consisted of 4,249 targeted-therapy–related cAEs in patients with lung cancer meeting all predefined inclusion criteria (Figure 6).

**Figure 6.**
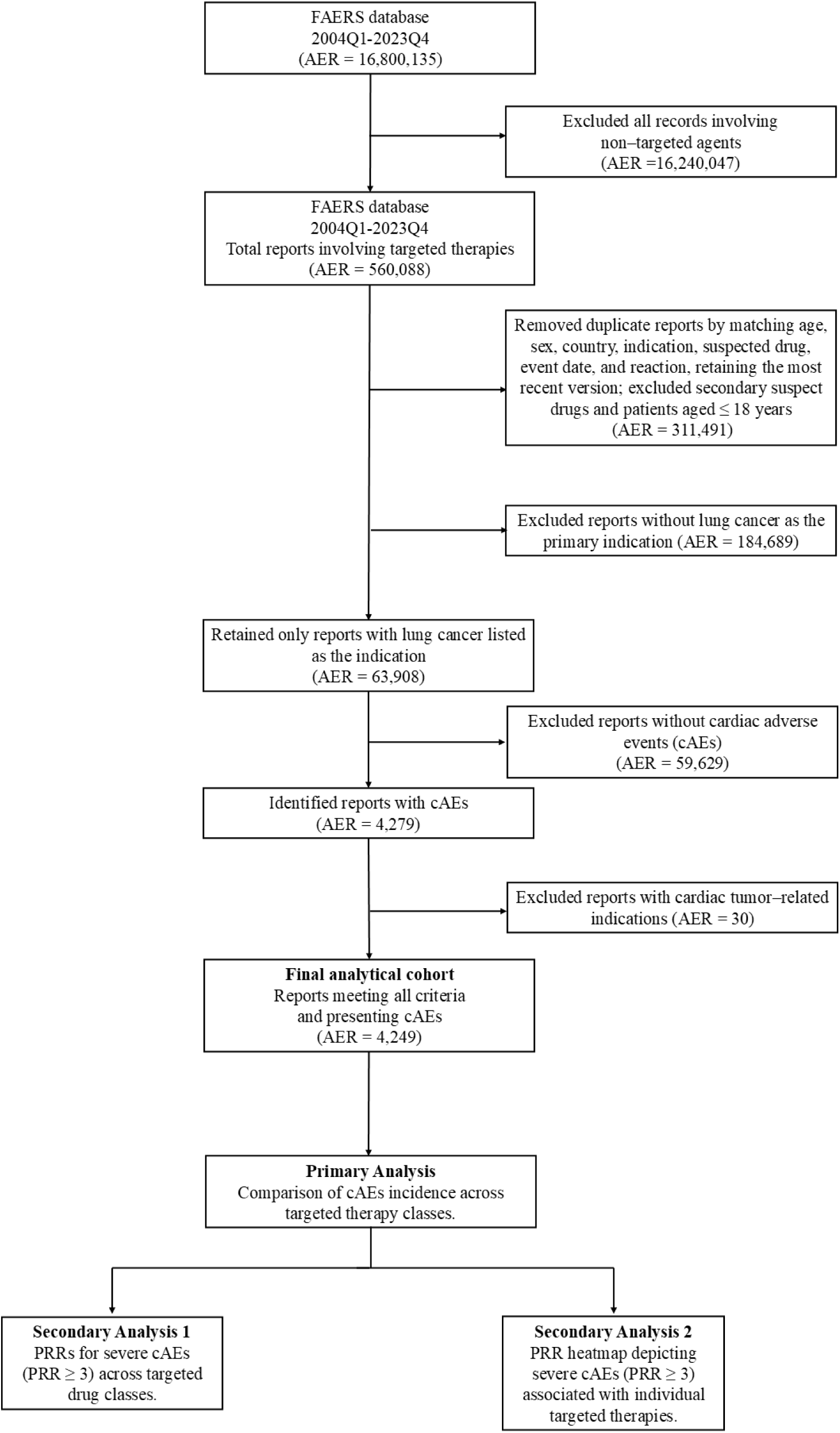
Flowchart of study analysis for cardiac adverse events in targeted drug therapy using FAERS database. This flowchart outlines the study’s analytical process in assessing cardiac adverse events (cAEs) related to targeted drug therapies using the FAERS database (2004Q1–2023Q4). The analysis includes three steps: (1) comparison of cAE incidence across different targeted therapy classes, (2) assessment of the PRR for severe cAEs (PRR ≥ 3) across targeted drug classes, and (3) PRR heatmap visualization for severe cAEs (PRR ≥ 3) associated with specific targeted therapies in lung cancer patients. The final cohort includes 4,249 reports of severe cAEs after data cleaning, exclusion of non-lung cancer cases, and deduplication. **Abbreviations: AER, adverse event report; cAE, cardiac adverse event; FAERS, FDA Adverse Event Reporting System; PRR, proportional reporting ratio.**

In the final analytical cohort, the incidence of cAEs differed markedly across targeted therapy classes, as the primary analysis. In secondary analyses, we characterized the proportional reporting ratios (PRRs) for severe cAEs (PRR ≥ 3) across targeted therapy classes and visualized drug-specific PRR patterns using a heatmap.

### Cardiovascular safety profile of different drug targets in lung cancer treatment in FAERS database

#### Overall cAEs frequency by drug target

The frequency of cAEs differed substantially across therapeutic targets (Figure 7A). PD-1 inhibitors exhibited the highest incidence of cAEs, with 8.09% of PD-1–treated patients affected. This was followed by VEGF inhibitors (7.93%) and ALK inhibitors (6.98%). EGFR inhibitors showed a cAE incidence of 5.71%, PD-L1 inhibitors 6.34%, and HER2 inhibitors had the lowest incidence at 4.23%.

**Figure 7.**
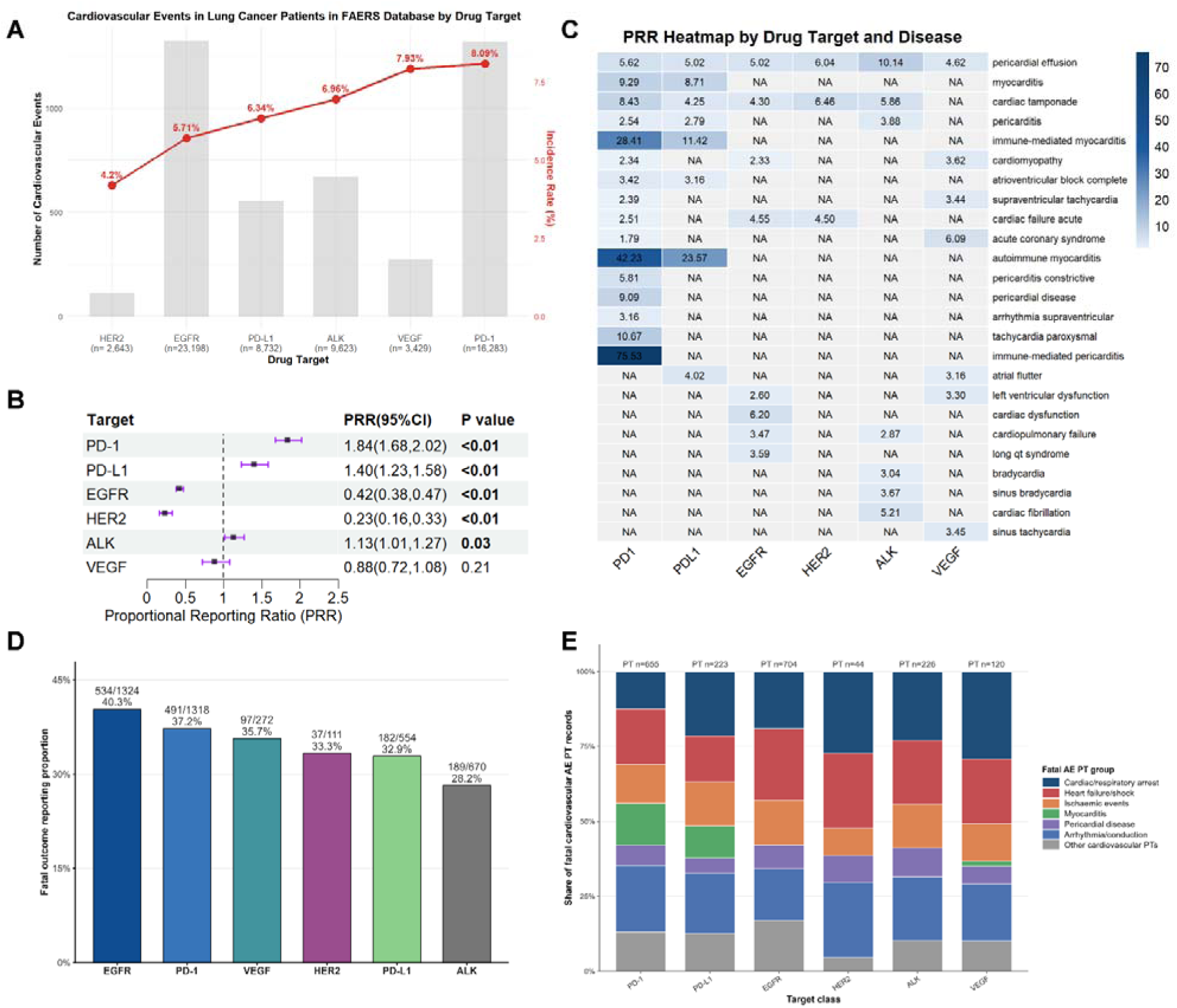
Cardiovascular safety profile of targeted therapies and immune checkpoint inhibitors in lung cancer patients: FAERS database analysis. (A) The line graph depicts the frequency distribution of cardiovascular adverse events (cAEs) across six drug target categories (HER2, EGFR, PD-1/L1, ALK, VEGF, and PD-1), with the percentage increase indicated above each data point. Cardiovascular event incidence rates (red line) and absolute event counts (gray bars) stratified by drug target in lung cancer patients. n represents the total number of AERs per drug target. (B) Forest plot displaying the proportional reporting ratio (PRR) with 95% confidence intervals (CI) for severe cAEs associated with each drug target. The vertical dashed line represents PRR = 1.0. (C) Heatmap illustrating the PRR values for specific severe cardiac adverse events (with ≥3 reported with darker blue indicating stronger disproportionality signals. NA indicates insufficient data (fewer than 3 cases) for PRR calculation. **Abbreviations:** ALK, anaplastic lymphoma kinase; cAE, cardiac adverse event; CI, confidence interval; EGFR, epidermal growth factor receptor; FAERS, FDA Adverse Event Reporting System; HER2, human epidermal growth factor receptor 2; NA, not available; PD-1, programmed cell death protein 1; PD-L1, programmed death-ligand 1; PRR, proportional reporting ratio; VEGF, vascular endothelial growth factor.

#### Disproportionality analysis of severe cAEs

Forest plot analysis demonstrated significant disproportionality signals for several drug targets (Figure 7B). PD-1 inhibitors exhibited the strongest association with severe cAEs, with a PRR of 1.84 (95% CI, 1.68–2.02, *P* < 0.01). PD-L1 inhibitors also showed a significant signal with a PRR of 1.40 (95% CI, 1.23–1.58, *P* < 0.01).

EGFR inhibitors were associated with a reduced cAE reporting signal (PRR 0.42; 95% CI, 0.38–0.47; *P* < 0.01), while HER2 inhibitors exhibited an even lower signal strength (PRR 0.23; 95% CI, 0.16–0.33; *P* < 0.01). ALK inhibitors showed a modest increased association with a PRR of 1.13 (95% CI, 1.01–1.27, *P* = 0.03), while VEGF inhibitors demonstrated no significant disproportionality with a PRR of 0.88 (95% CI, 0.72–1.08, *P* = 0.21).

#### Specific cAEs pattern across drug targets

The heatmap analysis revealed distinct patterns of specific cAEs associated with different drug targets (Figure 7C). Immune-mediated myocarditis showed the strongest disproportionality signal with PD-1 inhibitors (PRR = 28.41) and PD-L1 inhibitors (PRR = 11.42), highlighting the unique cardiotoxicity profile of immune checkpoint inhibitors. Autoimmune myocarditis exhibited an even higher disproportionality, with a PRR of 42.23 for PD-1 inhibitors and 23.57 for PD-L1 inhibitors. In addition, immune-mediated pericarditis demonstrated an exceptionally strong signal with PD-1 inhibitors (PRR = 75.53), further supporting the heightened susceptibility of immune-related cardiac complications under checkpoint blockade. Pericardial effusion was significantly associated with ALK inhibitors (PRR = 10.14), consistent with known class effects of this therapeutic category. HER2 inhibitors demonstrated elevated signals for cardiac failure (PRR = 2.27) and cardiac tamponade (PRR = 6.46). VEGF inhibitors showed notable associations with acute coronary syndrome (PRR = 6.09) and supraventricular tachycardia (PRR = 3.44). Autoimmune myocarditis was predominantly associated with PD-1 inhibitors (PRR = 23.57).

#### Fatal outcome reporting among targeted therapy–related cAEs

Among the final 4,249 target-class–expanded cAE reports, formal FAERS death outcome was recorded in 1,530 target-report combinations. The fatal outcome reporting proportion differed across target classes. EGFR inhibitor–related cAE reports showed the highest proportion of death outcome (534/1,324, 40.3%), followed by PD-1 inhibitors (491/1,318, 37.2%), VEGF inhibitors (97/272, 35.7%), HER2 inhibitors (37/111, 33.3%), PD-L1 inhibitors (182/554, 32.9%), and ALK inhibitors (189/670, 28.2%) (Figure 7D).

The composition of fatal cardiovascular AE PT records also varied by target class (Figure 7E). In EGFR inhibitor reports, fatal PT records were most frequently related to heart failure/shock (169/704, 24.0%), followed by cardiac/respiratory arrest (134/704, 19.0%) and arrhythmia/conduction events (121/704, 17.2%). In PD-1 inhibitor reports, arrhythmia/conduction events accounted for the largest share of fatal PT records (145/655, 22.1%), followed by heart failure/shock (121/655, 18.5%) and myocarditis (92/655, 14.0%). PD-L1 inhibitor reports showed a similar immune checkpoint inhibitor–related pattern, with cardiac/respiratory arrest (48/223, 21.5%), arrhythmia/conduction events (45/223, 20.2%), and myocarditis (24/223, 10.8%) contributing to fatal PT records. In VEGF inhibitor reports, cardiac/respiratory arrest (35/120, 29.2%) and heart failure/shock (26/120, 21.7%) were the dominant fatal PT categories. ALK inhibitor reports were mainly composed of cardiac/respiratory arrest (52/226, 23.0%), arrhythmia/conduction events (48/226, 21.2%), and heart failure/shock (48/226, 21.2%), whereas HER2 inhibitor estimates were based on fewer fatal PT records (PT n = 44) and should be interpreted cautiously.

## Discussion

Our study, leveraging a large real-world cohort of 2,427 lung cancer patients exposed to 82 targeted agents, provides one of the most comprehensive assessments to date of cardiotoxicity profiles across major therapeutic classes. Approximately 15.8% of patients experienced biomarker-defined cardiotoxicity. Among therapeutic classes, PD-1 inhibitors conferred the highest cardiotoxic risk, whereas EGFR inhibitors were associated with a comparatively lower incidence of cardiac events. The results further suggest that BNP and CK-MB, rather than TnI alone, may serve as sensitive early indicators of immune-related myocardial stress during PD-1 therapy. While previous studies have highlighted PD-1–related myocarditis (17), our data reveal a broader spectrum of subclinical myocardial stress, captured notably by BNP and CK-MB rather than troponin alone. Consistently, analyses of 4,249 targeted-therapy–related cardiac adverse event reports in the FAERS database showed that PD-1 inhibitors exhibited the strongest disproportionality signals for severe cardiotoxicity, whereas EGFR inhibitors demonstrated the lowest signal strength, reinforcing the class-specific risk patterns observed in our real-world cohort.

Clinically, PD-1–based immune checkpoint inhibitors are among the agents most frequently implicated in immune-mediated myocarditis and are associated with substantial morbidity and mortality despite their low absolute incidence (18). Immune checkpoint blockade can trigger systemic immune activation that extends to cardiac tissues (19). PD-1 signaling is essential for restraining autoreactive T-cell responses and preserving myocardial immune tolerance; both cardiomyocytes and cardiac-resident immune cells rely on PD-1/PD-L1 interactions to limit cytotoxic activity (19). Inhibition of this pathway removes a key peripheral tolerance mechanism, enabling T-cell infiltration, proinflammatory cytokine release, and direct myocyte injury (20). Experimental models further support this biology: PD-1–deficient mice spontaneously develop autoimmune myocarditis, underscoring its nonredundant role in cardiac homeostasis (21).

However, across the broad spectrum of targeted therapies, comparative risk estimates remain limited because most available evidence derives from case series or selected clinical-trial populations (22). Large real-world analyses are therefore needed to contextualize cardiotoxicity profiles across different therapeutic classes. Our study addresses this gap by systematically comparing cardiotoxicity across major targeted therapy categories and demonstrating that PD-1 inhibitors confer the highest overall cardiotoxic burden. Notably, the cardiotoxicity we identified is not restricted to clinically overt myocarditis; instead, it predominantly reflects a broader spectrum of subclinical or evolving myocardial stress captured by BNP and CK-MB—phenotypes that prior work has largely overlooked due to its narrow focus on fulminant myocarditis.

Although prior studies indicate that clinically overt immune checkpoint inhibitor (ICI)-associated myocarditis most commonly presents within the first 1–2 treatment cycles (23), our restricted cubic spline analysis revealed a nonlinear pattern with a steep rise in biomarker-defined cardiotoxicity up to approximately four administrations. This apparent shift likely reflects differences in case definition and patient populations. Unlike earlier reports focused on clinically adjudicated myocarditis—typically characterized by abrupt, early presentations (23)—our real-world cohort captures a broader spectrum of myocardial injury, including subclinical or attenuated phenotypes that may require multiple cycles of exposure before surpassing biomarker thresholds. The more heterogeneous clinical characteristics of routine oncology populations, including older age, multimorbidity, and polypharmacy, may also contribute to the delayed inflection in risk (24). Therefore, the cycle-dependent pattern observed in our study should not be interpreted as contradicting prior evidence but rather as complementary, highlighting that early-onset toxicity dominates overt myocarditis, whereas cumulative or subclinical injury may manifest over several administrations. This pattern is consistent with an early phase of immune activation reported in mechanistic studies, in which PD-1 blockade enables rapid T-cell activation and clonal expansion (25); however, direct evidence linking cycle number to clonal kinetics is limited, and the observed inflection point more likely reflects cumulative subclinical injury in real-world settings.

The longitudinal biomarker trajectories further clarify the temporal heterogeneity of cardiac injury. BNP and CK-MB showed significant exposure-related divergence, whereas TnI remained largely stable over time. These differences likely reflect distinct biological pathways and timing of myocardial stress. BNP, secreted by ventricular myocytes in response to wall stretch, is an early and sensitive indicator of hemodynamic overload or diastolic dysfunction, often preceding structural injury (26). CK-MB, a cytosolic enzyme released during myocyte membrane permeability changes, represents a transitional stage of subclinical injury (27). In contrast, TnI elevation signifies frank myocyte necrosis and tends to appear later and transiently during acute myocarditis episodes (28). Although clinically overt immune checkpoint inhibitor (ICI)-associated myocarditis is characterized by acute necrotic injury and marked troponin release, such cases represent only a small and highly selected subset of the overall cardiotoxicity spectrum captured in published case series (29). In routine practice—particularly when systematic biomarker surveillance is applied—many PD-1–related cardiac abnormalities manifest instead as mild or asymptomatic elevations in markers of myocardial stress or membrane permeability rather than fulminant necrosis (30). Therefore, in a real-world population enriched for low-grade or evolving myocardial stress rather than catastrophic necrotic injury, sustained BNP and CK-MB changes with minimal troponin release are pathophysiologically consistent. Consequently, integrating multiple biomarkers with complementary temporal sensitivity may improve early detection of PD-1-related cardiac effects (13).

## Limitations

Several limitations should be acknowledged. As a single-center retrospective analysis, residual confounding cannot be fully excluded (31). Cardiotoxicity was defined biochemically rather than by imaging, potentially overestimating mild or transient injury; however, biomarker-based definitions improve sensitivity for early stress detection and align with current cardio-oncology recommendations. Surveillance intensity differed between groups, which may introduce detection bias (32).

## Conclusions

- PD-1 inhibitors showed the strongest and most consistent cardiotoxicity signal across both the Renji real-world cohort and the FAERS database.
- The cardiotoxic risk of PD-1 therapy peaks around four treatment cycles.
- BNP and CK-MB trajectories capture early subclinical myocardial stress, whereas troponin remains stable, reflecting distinct temporal phases of cardiac injury.

## Data Availability Statement

All data produced in the present study are available upon reasonable request to the authors.

## Consent for publication

Not applicable.

## Corresponding Authors

Correspondence to Fan Wu, Meng Jiang and Jun Pu.

## Clinical trial number

Not applicable.

